# Assessing Health Systems-level Feasibility of Task-Sharing and -Shifting: Cross-sectional Study from Indian District Hospitals

**DOI:** 10.64898/2026.03.24.26349243

**Authors:** Siddhesh Zadey, Yash Jawale

## Abstract

**Introduction:** Low- and lower-middle-income countries face chronic deficits in specialist surgeons, particularly in rural and remote settings. Task-sharing and -shifting (TSS) to other health worker cadres is a remedy supported by effectiveness studies. However, policy adoption depends not only on effectiveness but also on feasibility at the health systems level, especially when personnel deficits co-occur across multiple cadres. We aimed to develop and apply an analytic approach to assess whether co-occurring workforce deficits constrain the feasibility of surgical TSS using a pan-Indian sample of district hospitals.

**Methods:** We conducted a cross-sectional descriptive analysis using administrative staffing data from the National Institution for Transforming India (NITI) Aayog assessment of 707 district hospitals for the 2017–18 financial year. Cadre-wise staffing ratios for specialist surgeons, general doctors, nurses, and paramedics were calculated using Indian Public Health Standards (IPHS) norms. A ratio <1 indicated a staffing deficit. We evaluated TSS feasibility for selected cadre pairs, with target and substituting cadres, using a rule-based categorization of relative staffing adequacy (not needed, possible, strained, unlikely, or not possible).

**Results:** Across 479 hospitals with deficits in specialist surgeons or general doctors included in the analysis. Staffing deficits were widespread across cadres. Only one hospital demonstrated TSS feasibility across all cadre pairs. For general doctors substituting specialist surgeons, TSS was possible in 1.5% of hospitals, strained in 9.0%, unlikely in 78.7%, and not possible in 10.9%. Similar patterns were observed for nurses substituting for specialist surgeons and for those substituting for general doctors. Feasibility was higher when paramedics substituted for general doctors or nurses, but co-occurring deficits were common.

**Conclusion:** Co-occurring workforce deficits substantially limit health systems-level feasibility of surgical task-sharing and -shifting in Indian district hospitals. Evaluation of TSS interventions should incorporate system-level workforce constraints in addition to evidence of effectiveness.

## Introduction

Low- and lower-middle-income countries, as well as rural and remote regions in high- and upper-middle-income countries, face chronic shortages of specialist surgeons.[1] Shifting surgical tasks to other available health workers, such as non-specialist/generalist physicians, nurses, and technicians, or sharing them among workers could help meet clinical needs in such cases. The last two decades have noted convergent evidence on the effectiveness and cost-effectiveness of surgical task-sharing and -shifting (TSS) interventions across tasks, cadres, settings, and outcomes.[2–8] The Lancet Commission on Global Surgery and the recent article in The Lancet, Surgical Health Policy 2025-35, noted the critical role of TSS in the medium term for addressing surgical workforce deficits.[9,10] Such evidence and arguments, while necessary, are not sufficient for policy action.

Several factors across different levels of the policymaking ecosystem influence action. One such factor is system-level feasibility. Consider a package of tasks (or a task) that should be shifted to or shared among health worker cadres. Now, assume that this package has been shown to be effective and cost-effective in an adequately powered, well-conducted randomized controlled trial in an appropriate setting. In line with evidence-based policymaking norms, the package would be scaled up for adoption across health facilities. However, whether such a scale-up works depends on the health system’s capabilities and constraints. In health systems facing chronic deficits across all health worker cadres, scaling up TSS might be challenging.[11–13] Tasks can be shifted to or shared among the cadres; however, this would increase the workload for everyone involved, leading to burnout. Hence, beyond evidence of a TSS intervention’s effectiveness, consideration of the health system-level feasibility is important.

Here, we present an analytic approach that demonstrates how personnel deficits in health worker cadres co-occur and that determines whether surgical TSS is feasible, using data from a pan-Indian sample of district hospitals.

## Methods

We used the report on ‘Best Practices in the performance of District Hospitals’ by the National Institution for Transforming India (NITI) Aayog, the premier governmental policy think tank.[14] The report presents an assessment conducted by the NITI Aayog in 2018-19. This assessment used the data from the Health Management Information System (HMIS) of 707 district hospitals for the 2017-18 financial year. The district hospitals are public-sector first-level surgical care hospitals that provide free/subsidized care in India.

We conducted a cross-sectional analysis of district hospital-level workforce capacity using administrative staffing data. For each district hospital, we used cadre-wise staffing ratios in accordance with the Indian Public Health Standards (IPHS) to assess the adequate availability of personnel across cadres. The IPHS staffing norms vary by hospital size, which in turn is characterized by the number of beds. **Table 1** provides the cadre-wise number of personnel required per IPHS norms for each group of district hospitals.

**Table 1:**
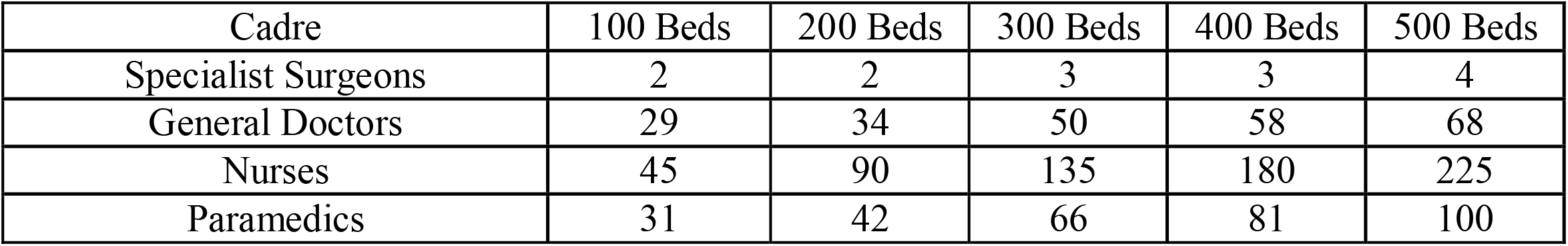
Indian Public Health Standards Staffing Norms for District Hospitals.

| Cadre | 100 Beds | 200 Beds | 300 Beds | 400 Beds | 500 Beds |
| --- | --- | --- | --- | --- | --- |
| Specialist Surgeons | 2 | 2 | 3 | 3 | 4 |
| General Doctors | 29 | 34 | 50 | 58 | 68 |
| Nurses | 45 | 90 | 135 | 180 | 225 |
| Paramedics | 31 | 42 | 66 | 81 | 100 |

Figure 1. provides a schematic summary of our analytic approach. Each ratio was defined as the observed number of cadre-specific personnel divided by the IPHS-recommended number for that hospital. The NITI Aayog report provided ratios for general doctors, nurses, and paramedical staff (paramedics). The surgeon staffing ratio was then calculated as the observed number of specialized surgeons divided by the hospital-level specific IPHS norm. We interpreted a ratio < 1 as a staffing deficit, whereas a ratio ≥ 1 indicated that IPHS norms were met or exceeded. We excluded hospitals with missing information on staffing ratios.

**Figure 1:**
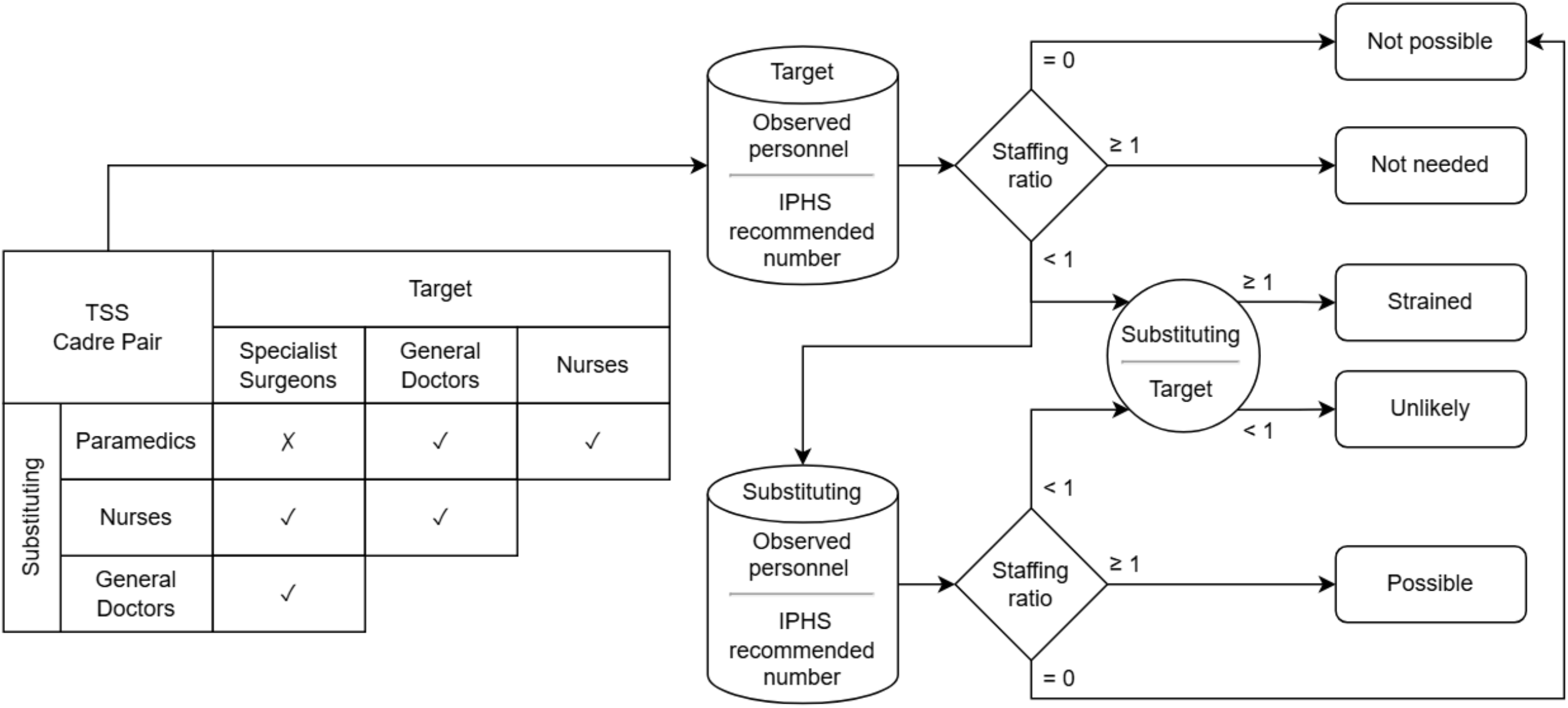
Schematic for the parsimonious rule-based analytic approach for task-sharing and - shifting feasibility at the health system-level. Matrix notes the substituting-target cadre pairs included in the analysis. Cylinders denote the data for observed personnel of a cadre and the recommended number based on the IPHS norm for a given hospital. Rhombus note the conditions based on staffing ratio (= 0, < 1, ≥ 1) and circle notes the condition based on the ratio of the substituting to target cadre (< 1, ≥ 1).

To assess TSS feasibility, we first identified hospitals with a deficit (ratio < 1) in cadres routinely trained to perform surgeries, i.e., specialist surgeons with postgraduate training and general doctors with graduate training. Next, we assumed that the most parsimonious model of TSS would be one in which certain tasks are shifted from one cadre (the target cadre) to another (substituting cadre), or shared by personnel of two cadres.

We considered the following cadre pairs based on existing TSS literature and our knowledge in India: general doctors substituting for (→) specialist surgeons, nurses → general doctors, nurses → specialist surgeons, paramedics → general doctors, and paramedics → nurses. We did not consider certain pairs for whom the shifting and sharing of tasks are implausible, e.g., paramedics → specialist surgeons. For each hospital and each cadre pair, we assigned one of the five TSS feasibility categories using a rule-based classification based on relative staffing adequacy: a) TSS Not Needed: The target cadre met the IPHS norm (ratio ≥ 1), b) Possible: The substituting cadre met IPHS norm (ratio ≥ 1) and had staffing levels equal to or exceeding the target cadre, c) Strained: The substituting cadre was below IPHS norms (ratio < 1) but had staffing levels equal to or exceeding the target cadre, d) Unlikely: The substituting cadre was below IPHS norms and had staffing levels below the target cadre, e) Not possible: Either the target or the substituting cadre had no personnel present (ratio = 0). It is evident why the absence of substitute cadre personnel would render TSS impossible. We assumed that surgical TSS would require some form of guidance or supervision from the personnel in the target cadre. Hence, a complete absence of target cadre personnel would not be an ideal setup for successful and sustained TSS. E.g., if a district hospital has no specialist surgeons, there is no one to train general doctors and nurses; hence, making surgical TSS impossible. Overall, the above categories reflect increasing constraints on the feasibility of redistributing clinical tasks under existing workforce availability **(Figure 1)**.

We calculated the frequency and percentage distributions of hospitals across TSS categories (Not needed, Possible, Strained, Unlikely, Not possible) for each cadre pair to facilitate comparison of feasibility patterns. The analysis was descriptive and did not involve statistical inference. We conducted all data processing and analyses in R (Version 4.4.3).

## Results

Of the 707 district hospitals in the source data, 52 were excluded due to missing staffing information. Further, 479 hospitals exhibited a deficit in either specialist surgeons or general doctors (ratio < 1) and were included in the TSS feasibility assessment. Within the analytic sample, deficits were widespread across cadres. One hundred and thirty-one district hospitals had deficits in specialist surgeons, and 470 had deficits in general doctors. Deficits in staffing of nurses and paramedics affected 447 and 228 hospitals, respectively.

Only one district hospital in Sangrur, Punjab, showed that TSS was ‘possible’ for all cadre pairs, indicating that health system-wide TSS feasibility was rare. Below, we describe results by cadre pairs. **Figure 2** provides a concise summary of these findings.

**Figure 2:**
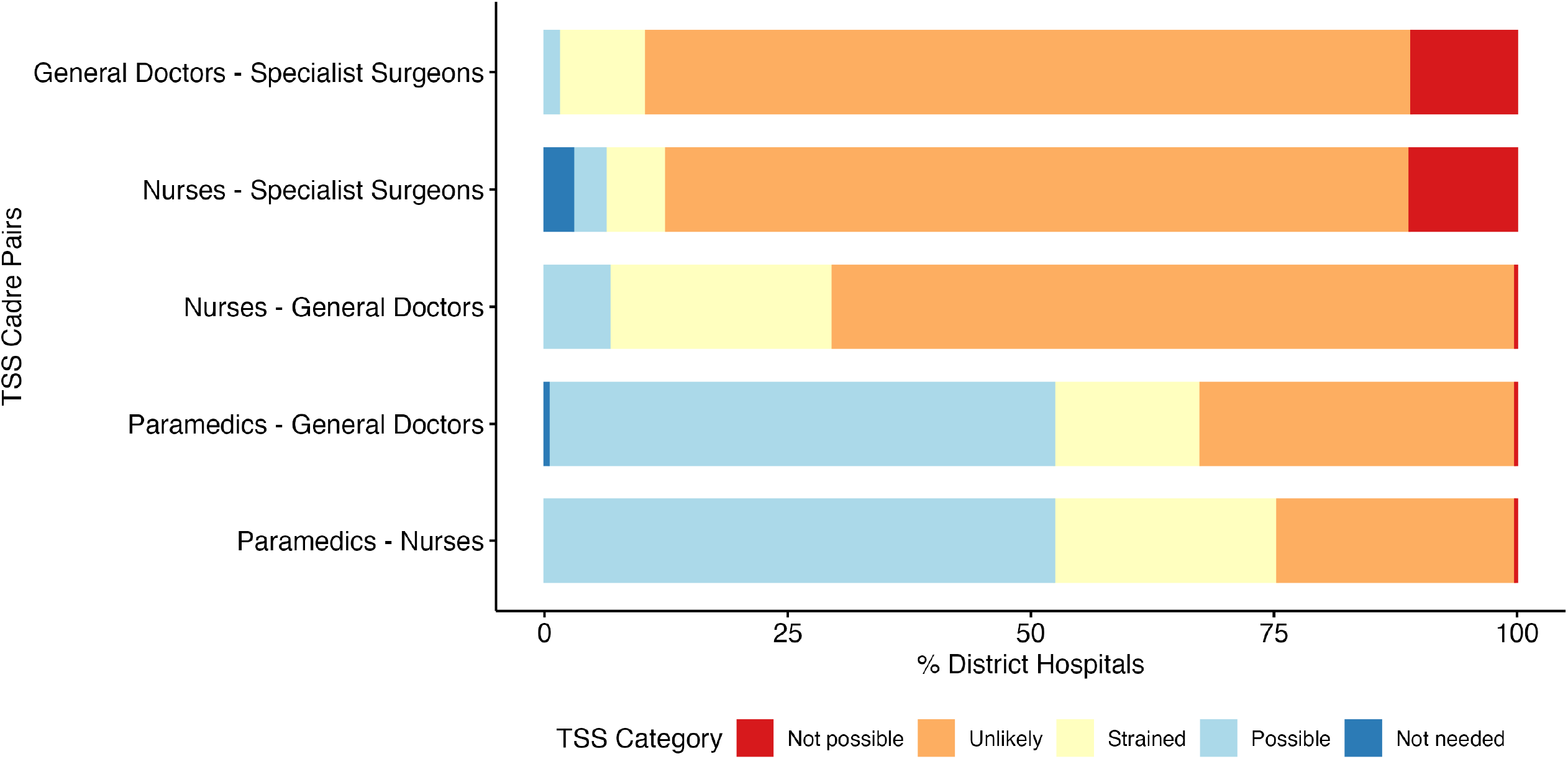
Feasibility of task-shifting and -sharing for different cadre pairs at district hospitals in India.

For the general doctors-specialist surgeons cadre pair, TSS was categorized as ‘possible’ in 7 (1.5%) hospitals. TSS was ‘unlikely’ for 377 (78.7%) hospitals, while 43 (9.0%) hospitals had potential for TSS that would have been ‘strained’. Per our analytic approach, general doctors substituting in for specialist surgeons for some tasks was not possible in 52 (10.9%) hospitals due to the absence of specialist surgeons.

For the nurses-specialist surgeons cadre pair, TSS was ‘not needed’ at 14 (2.9%) hospitals, because the IPHS norm was met for specialist surgeon staffing. TSS was ‘possible’ at 16 (3.3%) hospitals and would have been ‘strained’ at 30 (6.3%) hospitals. TSS was ‘unlikely’ at the majority of hospitals (366; 76.4%). TSS was ‘not possible’ at 53 (11.1%) hospitals due to the absence of specialist surgeons or nurses.

For the nurses-general doctors cadre pair, TSS was ‘possible’ at 32 hospitals (6.7%), and ‘strained’ at 110 hospitals (23.0%). TSS was ‘unlikely’ at the majority of hospitals (336; 70.2%). TSS was ‘not possible’ at one hospital due to the absence of nurses.

For the paramedics-general doctors cadre pair, TSS was ‘not needed’ at 2 hospitals where general doctors met the IPHS norm. TSS was ‘possible’ at 249 (52.0%) hospitals and would have been ‘strained’ at 72 (15.0%) hospitals. TSS was ‘unlikely’ at 155 (32.4%) hospitals. TSS was ‘not possible’ at one hospital due to the absence of paramedics.

For the paramedics-nurses cadre pair, TSS was ‘possible’ at 251 (52.4%) hospitals and would have been ‘strained’ at 110 (23.0%) hospitals. TSS was ‘unlikely’ at 117 (24.4%) hospitals. TSS was ‘not possible’ at one hospital due to the absence of paramedics and nurses.

## Discussion

Overall, this cross-sectional pan-India analysis demonstrates pervasive staffing deficits and the limited feasibility of task-sharing and -shifting in most district hospitals, especially for specialist surgeons and general doctors.

A couple of related observations are noteworthy. First, we underscore that adequate personnel in the substituting cadre are a prerequisite. E.g., interventions that shift tasks from specialist surgeons to general doctors or nurses are most feasible where general doctors and nurses are adequately present and have to absorb additional responsibilities. Second, co-occurring workforce deficits in health facilities are a health systems issue governed by multi-level, interrelated factors, including budget allocations, hiring policies, incentives, and work environments. Hence, it should be treated as such. E.g., even if one introduces a large number of surgical specialists into the health system, deficits in other cadres (e.g., nurses and general doctors) may limit the practical ability to redistribute tasks without overburdening the remaining workforce. Solving such issues requires appropriate systems thinking.[15,16] Third, beyond systems-level feasibility, the success of TSS interventions also relies on broader organisational and regulatory environments. E.g., a feasible TSS intervention may still not work in practice if it is opposed by established professional surgical organizations or by rigid regulatory barriers.[12,17,18]

This study has several strengths. First, we develop and apply a parsimonious, rule-based, interpretable analytic approach to assess the feasibility of TSS. Our approach can be adapted to other settings where such health workforce staffing data and contextually relevant norms. We circumvent reliance on complex statistical modelling and transparently present a small set of assumptions that are justifiable in settings with heterogeneous data availability and quality. Second, as an application of our analytic approach, we leverage a large, facility-level dataset from a lower-middle-income country context, enabling a granular assessment of workforce availability at the district hospital level, an analytic scale that is rarely available in health systems research. Third, by anchoring workforce staffing adequacy to the IPHS norms, the analysis aligns with nationally recognized policy benchmarks. This strengthens the relevance and potential translational value of the findings for workforce planning and health system strengthening in India.

This study also has important limitations. First, our approach does not explicitly account for the complexities of surgical teams, including task interdependencies, supervision requirements, and differences in skill mix within and between cadres. These may impose further constraints on the real-world TSS feasibility beyond what staffing ratios captured. Second, the approach does not capture variation in clinical environments, such as case mix, infrastructure readiness, or informal adaptations to service delivery governed by local contextual practices. E.g., there could be instances in which paramedics might be able to perform tasks typically performed by nurses due to contextual norms. In such instances, paramedics → specialist surgeons, TSS, which we excluded in the current analysis, could be considered. Such variations may influence whether TSS is feasible or safe in reality, beyond the parsimonious structure considered in the study. Third, although IPHS norms provide a useful reference from an Indian policymaking perspective, they have previously been noted to underestimate true workforce requirements.[11] This implies that workforce staffing deficits and constraints on TSS may be more severe than reported.

## Conclusion

Our transparent, interpretable, and reproducible analytic approach reveals how health-systems-level factors, such as the co-occurrence of workforce deficits, can limit the feasibility of adopting and sustainably running task-sharing and -shifting interventions. Future studies can extend the approach to account for more complex surgical TSS scenarios across varied settings. Regardless, accounting for and addressing health systems-level issues is critical to the sustained scale-up of TSS across diverse global settings.

## Data Availability

All data produced in the present study are available upon reasonable request to the authors.

## Acknowledgements

We would like to acknowledge Madhurima Vuddemarry and Anveshi Nayan, from ASAR, for assistance with initial data management.

## Notes

**Conflicts of interest statement:** Siddhesh Zadey serves as the co-founding director of the Association for Socially Applicable Research (ASAR), Chair of the Asia Working Group of the G4 Alliance, Fellow at the Lancet Commission on a Citizen-Centred Health System for India, and Drafting Committee Member for the Maharashtra State Mental Health Policy. The other authors declare no conflicts of interest.

### Competing Interest Statement

Siddhesh Zadey serves as the cofounding director of the Association for Socially Applicable Research (ASAR), Chair of the Asia Working Group of the G4 Alliance, Fellow at the Lancet Commission on a Citizen Centred Health System for India, and Drafting Committee Member for the Maharashtra State Mental Health Policy. The other authors declare no conflicts of interest.

### Funding Statement

This study did not receive any funding.

### Author Declarations

This article uses publicly available hospital-level aggregate data. It does not include human participant or animal subject data. Hence, approval from an ethics committee/institutional review board was not considered required.

## Bibliography

1. Holmer H, Lantz A, Kunjumen T, Finlayson S, Hoyler M, Siyam A, et al. Global distribution of surgeons, anaesthesiologists, and obstetricians. Lancet Glob Health. 2015 Apr 27;3 Suppl 2:Supplementary Appendix Table S4.

2. Ryan I, Shah KV, Barrero CE, Uamunovandu T, Ilbawi A, Swanson J. Task Shifting and Task Sharing to Strengthen the Surgical Workforce in Sub-Saharan Africa: A Systematic Review of the Existing Literature. World J Surg. 2023 Dec;47(12):3070–80.

3. Ashengo T, Skeels A, Hurwitz EJH, Thuo E, Sanghvi H. Bridging the human resource gap in surgical and anesthesia care in low-resource countries: a review of the task sharing literature. Hum Resour Health. 2017 Nov 7;15(1):77.

4. Falk R, Taylor R, Kornelsen J, Virk R. Surgical Task-Sharing to Non-specialist Physicians in Low-Resource Settings Globally: A Systematic Review of the Literature. World J Surg. 2020 May;44(5):1368–86.

5. Federspiel F, Mukhopadhyay S, Milsom PJ, Scott JW, Riesel JN, Meara JG. Global surgical, obstetric, and anesthetic task shifting: A systematic literature review. Surgery. 2018 Sep;164(3):553–8.

6. Bognini MS, Oko CI, Kebede MA, Ifeanyichi MI, Singh D, Hargest R, et al. Assessing the impact of anaesthetic and surgical task-shifting globally: a systematic literature review. Health Policy Plan. 2023 Sep 18;38(8):960–94.

7. Oko CI, Ali B, Monahan M, Aborode AT, Okon J, Ayomoh F, et al. Economics of Task-Shifting in Surgery: A Systematic Review. Health Sci Rep. 2025 Sep 3;8(9):e71198.

8. Hoyler M, Hagander L, Gillies R, Riviello R, Chu K, Bergström S, et al. Surgical care by non-surgeons in low-income and middle-income countries: a systematic review. Lancet. 2015 Apr 27;385 Suppl 2:S42.

9. Meara JG, Leather AJM, Hagander L, Alkire BC, Alonso N, Ameh EA, et al. Global Surgery 2030: evidence and solutions for achieving health, welfare, and economic development. Lancet. 2015 Aug 8;386(9993):569–624.

10. Nepogodiev D, Picciochi M, Ademuyiwa A, Adisa A, Agbeko AE, Aguilera M-L, et al. Surgical health policy 2025-35: strengthening essential services for tomorrow’s needs. Lancet. 2025 Jul 14;

11. Nair A, Jawale Y, Dubey SR, Dharmadhikari S, Zadey S. Workforce problems at rural public health-centres in India: a WISN retrospective analysis and national-level modelling study. Hum Resour Health. 2022 Jan 28;19(Suppl 1):147.

12. Zadey S. Task sharing and shifting: Can we move beyond effectiveness? [Internet]. Health Systems Global (HSG). 2025 [cited 2026 Feb 19]. Available from: https://healthsystemsglobal.org/news/task-sharing-and-shifting-can-we-move-beyond-effectiveness/

13. Gilles I, Le Saux C, Zuercher E, Jubin J, Roth L, Bachmann AO, et al. Work experiences of healthcare professionals in a shortage context: analysis of open-ended comments in a Swiss cohort (SCOHPICA). BMC Health Serv Res. 2025 Apr 9;25(1):520.

14. Sarwal R, Iyer V, Kalal S. Best practices in the performance of District Hospitals in India. 2021 Oct 7;

15. Don de Savigny and Taghreed Adam (Eds). Systems thinking for health systems strengthening. WHO; 2009.

16. Atun R, Menabde N. Health systems and systems thinking. Health systems and the challenge of …. 2008;

17. van Heemskerken P, Broekhuizen H, Gajewski J, Brugha R, Bijlmakers L. Barriers to surgery performed by non-physician clinicians in sub-Saharan Africa-a scoping review. Hum Resour Health. 2020 Jul 17;18(1):51.

18. Shah NM, Afsar AP, Zadey S. Rethinking Global Surgery Research: Role of Mixed-Methods Studies. The American Journal of Surgery. 2026 Jan;116831.

